# Children’s and Parents’ Perspectives on Universal Free School Meals in Wales: A Mixed Methods Study on Health, Wellbeing and Barriers to Uptake

**DOI:** 10.64898/2026.03.02.26347394

**Authors:** Amy Locke, Michaela James, Sinead Brophy

## Abstract

**Background:** Universal Free School Meals (UFSM) were introduced by the Welsh Government in 2022 to provide every primary school child (aged 4-11) with a free meal at lunch time by 2024, aiming to improve nutrition and reduce inequality. While evidence suggests UFSM can support dietary quality and social inclusion; uptake remains inconsistent, and concerns have been raised regarding meal quality and portion sizes.

**Aims/Objectives:** This study explored the perceptions of children and parents regarding the rollout of UFSM in Wales, focusing on perceived health, wellbeing and social impacts whilst also identifying factors influencing non-uptake.

**Methods:** A mixed-methods study was conducted, combining qualitative focus groups with 56 children in year 6 (aged 10-11) across eight primary schools in Wales. A cross-sectional survey was also completed by 410 parents from 110 Welsh primary schools. Qualitative data from focus groups and open-ended survey questions were analysed thematically using Braun and Clarke’s framework, whilst closed-ended survey items were analysed descriptively to complement and contextualise qualitative findings.

**Results:** Three themes emerged from the children’s data. (1) The Food Experience, (2) The Social Value of Lunchtime, and (3) Fuel for Learning and Feeling Good. Children valued the social and emotional aspects of mealtimes but reported mixed experiences with food quality, variety, and portion sizes. Parents similarly expressed concerns about meal nutritional quality but also highlighted the policy’s benefits in reducing financial strain, stress, and daily time pressures.

**Conclusions:** UFSM is widely supported for promoting inclusion and access to food. Nevertheless, improving meal quality, portion sizes, and menu diversity is essential to sustain participation and maximise the policy’s long-term health and equity benefits.

## Introduction

Children’s diets play a crucial role in their health, wellbeing, and overall development [1]. A balanced diet of fruit and vegetables, wholegrains, protein-rich foods and dairy support children’s physical growth, cognitive development, and emotional welling [2, 3]. In the UK, dietary guidance for primary school children recommends a varied diet. In line with the Eatwell Guide [4], children should consume at least five portions of fruit and vegetables per day, starchy carbohydrates and limited consumption of foods high in saturated fat, salt and free sugars [5].

School meals, that is meals provided at a designated lunch time slot, make a substantial contribution to children’s nutritional intake, providing one third of their daily energy and micronutrient needs [6,7]. Research has shown that school meals are also typically a healthier option than packed lunches from home, offering greater variety and higher nutritional quality [8]. Evidence from longitudinal studies suggest that children’s eating habits often continue into adolescence and adulthood, highlighting the importance of establishing healthy eating behaviours in early childhood [9]. Therefore, improving uptake of UFSM may support access to healthier food options and reinforce eating habits [10].

In recognition of the role that nutrition plays in children’s health and development, the Welsh Government introduced Universal Free School Meals (UFSM) from 2022 [11]. The policy committed to providing every primary school child in Wales with a free meal, regardless of household income, by 2024. This policy introduction replaces the previous means-tested model in which eligibility depended on household income [12]. To support implementation, the Welsh Government committed dedicated funding to LA’s and schools to expand catering capacity, upgrade kitchen facilities and support staffing requirements associated with the phased-roll out [13], allowing schools to adapt to increased school meal demands.

According to a report by NESTA [14], despite the introduction of this policy, 70,000 (30%) of children are still opting to bring a packed lunch to school. Reasons for this have been explored in a recent survey conducted by the Children’s Commissioner for Wales (2024) [15]. The survey, including 490 respondents aged between 7-18, noted that only 19% of children reported feeling full after their school meal. Respondents also reported inconsistent access to fruit (22%) and vegetables (24%). Qualitative responses frequently mentioned inadequate portion sizes, with older children being served the same amount as younger children. The report concludes that while the UFSM policy is welcome, its effectiveness is undermined if meals fail to meet children’s nutritional needs.

In addition to the nutritional impact of UFSM, little is known about the social and wellbeing impacts of the policy. Emerging evidence suggests a range of social; emotional and wellbeing benefits associated with UFSM provision. For example, a recent scoping review by Locke et al. [16] which mapped international evidence of targeted and universal school meals provision, found that UFSM reduced stigma which is normally associated with means-tested provision. It also saw an increase in children’s sense of belonging through improved social inclusion. The review also noted that through shared mealtimes and dining experiences, children’s communication, cooperation and turn-taking also improved over time.

Further, work from Norway has demonstrated that UFSM helps remove visible markers of poverty, improving peer-relationships [17]. Similarly, a U.S study reported reductions in bullying and improvements in the school social climate following implementation of UFSM [18]. These findings suggest broader psychosocial benefits of UFSM and highlight school mealtimes as an opportunity to promote children’s social development, peer connections as well as emotional wellbeing outcomes.

Much of this research comes from countries with different school food systems, nutritional regulations and cultural expectations. Therefore, it is unclear if similar experiences and outcomes would be seen in Wales, particularly given the rapid rollout of UFSM across diverse Welsh primary schools. Locally grounded research is needed to understand how children and families are experiencing the policy in practice across Wales.

Despite growing evidence that UFSM provision can improve children’s dietary quality, reduce stigma and enhance social inclusion, gaps remain in understanding how these policies are experienced. In Wales, where UFSM has recently been implemented, uptake varies across schools and concerns remain around meal quality and portion sizes. Limited research has explored how children and families perceive the policy’s implementation or how UFSM influences daily routines, wellbeing, and social dynamics within schools. Whilst existing data such as uptake level [14] provide useful insights, they offer limited understanding of how UFSM is experienced in daily school life. Qualitative perspectives are needed to understand the social, emotional and contextual factors shaping children and family’s engagement with the provision. Therefore, this study aims to explore the perceptions of children and parents regarding the rollout of UFSM in Wales, with a specific focus on perceived health and wellbeing impacts and the factors influencing uptake.

## Methods

### Design

A mixed-methods design was employed to explore the experiences and perspectives of children and parents on the implementation of UFSM in Wales. Qualitative data was collected through focus groups with children to capture in-depth accounts of mealtime experiences. Additionally, a cross-sectional online survey for parents was conducted, which included both closed-ended and open-ended questions.

### Participants

#### Children

Twenty primary schools across Wales were invited by phone or email between October 2024 and May 2025. Schools were selected to capture a range of perspectives from different geographical areas of Wales to reflect potential variation in how UFSM was being implemented across local authorities (LA). Schools from multiple regions (Swansea, Llanelli, Cardiff, Aberdare, and Wrexham) were invited to fully represent the geography of Wales. A total of eight (40% of those invited) primary schools agreed to participate. Each school hosted one in-person focus group conducted between January 2025 and Junes 2025, with 6-8 children from year 6, resulting in a total of 56 participating children. These focus groups took place in the participating school, as part of the school day. Year 6 pupils were selected because they were considered developmentally able to participate confidently in group discussions.

Participating schools included a mix of urban and rural settings across several LAs. To provide contextual information about participating schools, publicly available school-level data was used, including school size, the proportion of pupils eligible for means-tested FSM prior to UFSM implementation based on publicly available information from my local school [19]. Area-level deprivation based on the Welsh Index of Multiple Deprivation (WIMD) 2019 [20] is also provided. WIMD scores were derived from the school catchment area and reflect area-level deprivation rather than the socio-economic circumstances of individual pupils. These data are presented in Table 1.

**Table 1.** Characteristics of participating primary schools and focus groups (n = 8 schools)

| **School** | **School size (No of pupils as of 2025)** | **% FSM eligible (pre-UFSM)** | **WIMD quintile** | **Focus group size** |
| --- | --- | --- | --- | --- |
| School A | 197 | 4.8% | 5 | 8 |
| School B | 461 | 26.5% | 1 | 6 |
| School C | 320 | 23.3% | 3 | 9 |
| School D | 166 | 31.7% | 2 | 8 |
| School E | 402 | 2.5% | 5 | 6 |
| School F | 204 | 47.7% | 2 | 5 |
| School G | 281 | 17.1% | 5 | 6 |
| School H | 220 | 19.8% | 3 | 8 |

#### Parents and Guardians

Parents and guardians were recruited through the HAPPEN (Health and Attainment of Pupils in Primary Education) school cohort, a pan-Wales collaboration that aims to empower primary schools to better support their children’s health, wellbeing and education [21,22,23]. In this study, schools shared the survey link with parents and guardians on behalf of the research team. The survey reached parents from at least 110 primary schools across Wales, indicating wide geographical coverage. The online survey was open from January to July 2025 and received 415 responses. 5 responses were removed during cleaning due to clicking no on the consent question, leaving 410 responses for analysis. Eligibility required participants to have at least one child attending a Welsh primary school.

### Data collection and processing

Focus group sessions took place during school hours in a private classroom to ensure confidentiality and comfort. Consent forms were sent home to parents prior to participation. Written consent was obtained from both parents and children. Children’s assent was verbally re-affirmed at the beginning of each session and monitored throughout, with researchers reminding pupils of their right to pause, skip questions, or withdraw at any time. A semi-structured discussion guide including 11 open-ended questions was used to explore pupils’ experiences. The topic guide was developed by lead researcher (AL) and reviewed for clarity; it was piloted with a small group of children at a local primary school to ensure age-appropriate wording. Questions included: ‘How do you feel after eating a school meal compared to when you eat a packed lunch?’ and ‘If you could create your own school lunch menu, what would it include?’ (See supplementary (S1) for full list of questions). Each focus group lasted approximately 30 minutes and was facilitated by two researchers (AL and MJ). Both facilitators had prior experience in conducting qualitative research with children. Focus groups were audio-recorded and transcribed within 24 hours by AL. The audio recording was deleted after transcription.

A cross-sectional online survey was created using Microsoft Forms to capture parent’s perspectives of UFSM. Questions combined Likert-scale items, multiple-choice and open-ended questions. Likert items asked parents their agreements with statements such as *“Free school meals reduce child hunger”*. Multiple choice questions explored frequency and patterns of uptake ((e.g., *“How often does your child eat school meals versus bringing food from home?”*) and open-ended questions invited more detailed views on perceived changes to children’s eating habits, wellbeing and attitudes toward healthy eating. Closed ended questions were included to contextualise parents qualitative accounts, rather than to test hypothesis or infer causal relationships. The full list of survey questions is provided in Supplementary Materials (S2). Parents completed the survey in their own time on their personal devices, allowing for flexible participation. Completion time varied, but most parents finished the survey within 8–10 minutes. Responses were exported into Microsoft Excel for cleaning and organisation prior to analysis.

### Analysis

Qualitative data was analysed using inductive thematic analysis based on Braun and Clarke’s six-phase framework [24, 25]. A within-group thematic analysis was conducted for each participant group (children, parents) to identify patterns and experiences specific to each group. Initial codes were generated line by line to capture meaningful features. Initial codes were then collated into themes. AL conducted the primary coding of all transcripts, and MJ independently reviewed a subset of the coded data to check consistency and enhance reliability. NVivo [26] software was used to manage and organise all qualitative data, including both children’s focus group transcripts and parents’ open-ended survey responses.

Closed-ended survey responses were analysed descriptively using IBM SPSS Statistics [27] to summarise percentages of responses. These quantitative findings were used to contextualise and support the qualitative themes in a complementary descriptive data allowing survey patterns (e.g., concerns about portion size) to be compared with qualitative perspectives provided by parents.

## Results

A total of 56 children across eight focus groups and 410 parents participated from across Wales. Participating schools varied in size and geographical context (urban and rural), although socio-economic characteristics were not collected. Five themes were identified across both focus groups and parent survey. Together, these themes illustrate how children and parents experience UFSM and what shapes mealtime satisfaction and participation.

Themes are presented below with illustrative quotations from children’s focus groups and parents’ survey responses. The complete themes table is provided in Supplementary (S3).

### Children’s Perspectives

Three themes were identified from the children’s focus groups: (1) The Food Experience, (2) The Social Value of Lunchtime, and (3) Fuel for Learning and Feeling Good.

### Theme One: The Food Experience

Across focus groups, children described how the quality, portion size, and variety of meals shaped their experiences of UFSM. While some pupils reported enjoying meals, such as the roast dinners, and trying new foods, others described dissatisfaction with taste, freshness, or repetitiveness. Positive experiences included trying meals they had not eaten before:

*“I like the fish fingers because I’d never tried them before and it caught my attention. It was really good.”* (Child 4, Focus group 5)

*“I like the pasta and the chicken dinner.”* (Child 2, Focus group 7) However, concerns about food quality and texture were common:

*“Most of the time I have it and it’s like all like soggy and raw. The outside of it is all like wet and the inside of it is like rock solid.”* (Child 5, Focus group 6)

*“I don’t really like the pizza because it’s like hard”* (Child 5, Focus group 4)

Portion sizes were frequently mentioned, although the satisfaction was different across LA’s:

*“On Friday when they do fish, they only give you like two fish fingers.”* (Child 3, Focus group 2)

Wanting more variety of food options was also common with vegetarian pupils expressing limited choice:

*“They’re quite bad now, especially for vegetarians. There isn’t enough choice, and the stuff doesn’t taste nice.”* (Child 2, Focus group 5)

Children’s accounts revealed that the quality, portion size and variety of meals differ across schools. Dissatisfaction with portion sizes, repetitive menus or unappealing food was frequently mentioned across groups, thus reducing enjoyment in the school meal overall.

These perceptions are contextualised by illustrative photographs of typical school meals provided during the study period (Figure 1).

**Figure 1:**
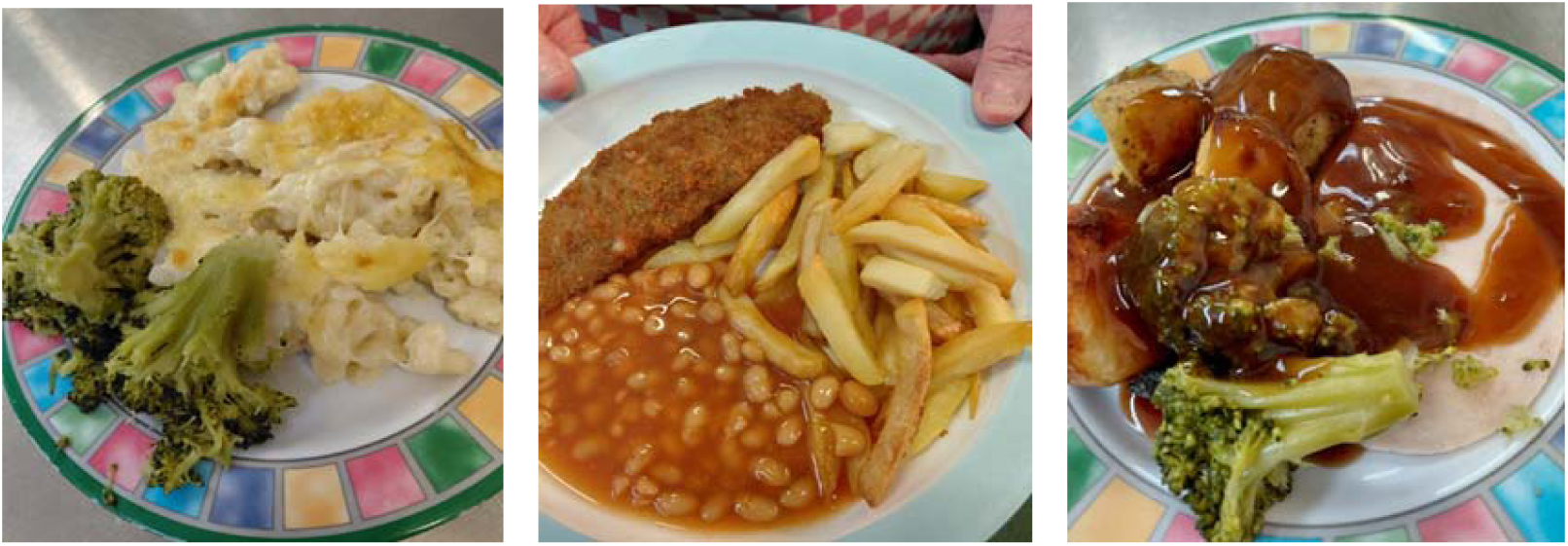
Example of a universal free school meals provided to a Welsh primary school pupil.

### Theme two: The Social Value of Lunchtime

Children emphasised the importance of lunchtime as a social experience providing a sense of belonging and a time to play. Several participants described improvements since the introduction of universal meals, as shared dining arrangements had created greater opportunities for inclusion:

*“It is better now because it used to be pack lunches on one table and dinners on another, so we didn’t get to sit with your friends if they were all packed lunch and you were dinners.”* (Child 5, Focus group 3)

However, some schools continued to have restrictions on seating:

*“We normally just go in and hope we can sit by our friends, because we don’t always get to choose.”* (Child 4, Focus group 3)

*“It’s not fair, because you want to sit by your friends. Like I sit by all the boys and it’s not fair.”* (Child 5, Focus group 3)

Lunchtime also played a role in facilitating social planning, and opportunity to play, extending the benefits of school meals beyond the canteen:

*“We make plans about what we are going to do after school or games we are going to play on.”* (Child 5, Focus group 3)

*“Then as soon as we’re done you could just bolt it for the field.”* (Child 7, Focus group 3)

*“So we could play!”* (Child 3, Focus group 3)

Overall, children’s reflections highlight the important social aspects of lunch time. While shared dining has promoted greater inclusion and interaction among peers, limited space and structured seating arrangements sometimes restricted opportunities for friendship and play. This emphasises that the success of UFSM is tied not only to what is served, but also to how lunchtime enables connection, autonomy, and a sense of belonging within the school day.

### Theme three: Fuel for Learning and Feeling Good

Children frequently linked eating a good meal with having energy, improved concentration, and a more positive mood.

*“School meals actually help me have energy through the day.”* (Child 5, Focus group 2)

*“If it looks nice, I will have a good day, but if it doesn’t look nice then I don’t want to eat it and I’ll have a bad day for the rest of the day.”* (Child 2, Focus group 6)

Pupils also noted that the school’s meals helped them concentrate on their learning:

*“When I have had a good meal, I won’t be thinking I want food because I’m hungry. I can think about the work.”* (Child 1, Focus group 4)

These accounts demonstrate how children’s emotional wellbeing was influenced by the lunchtime experience. Enjoyable, satisfying meals contributed not only to feeling energised and ready to learn but also to their overall happiness to continue the school day.

### Parents Perspectives

A total of 410 parents completed the cross-sectional Survey. Table 2 summarises the demographic characteristics of the survey sample.

**Table 2:**
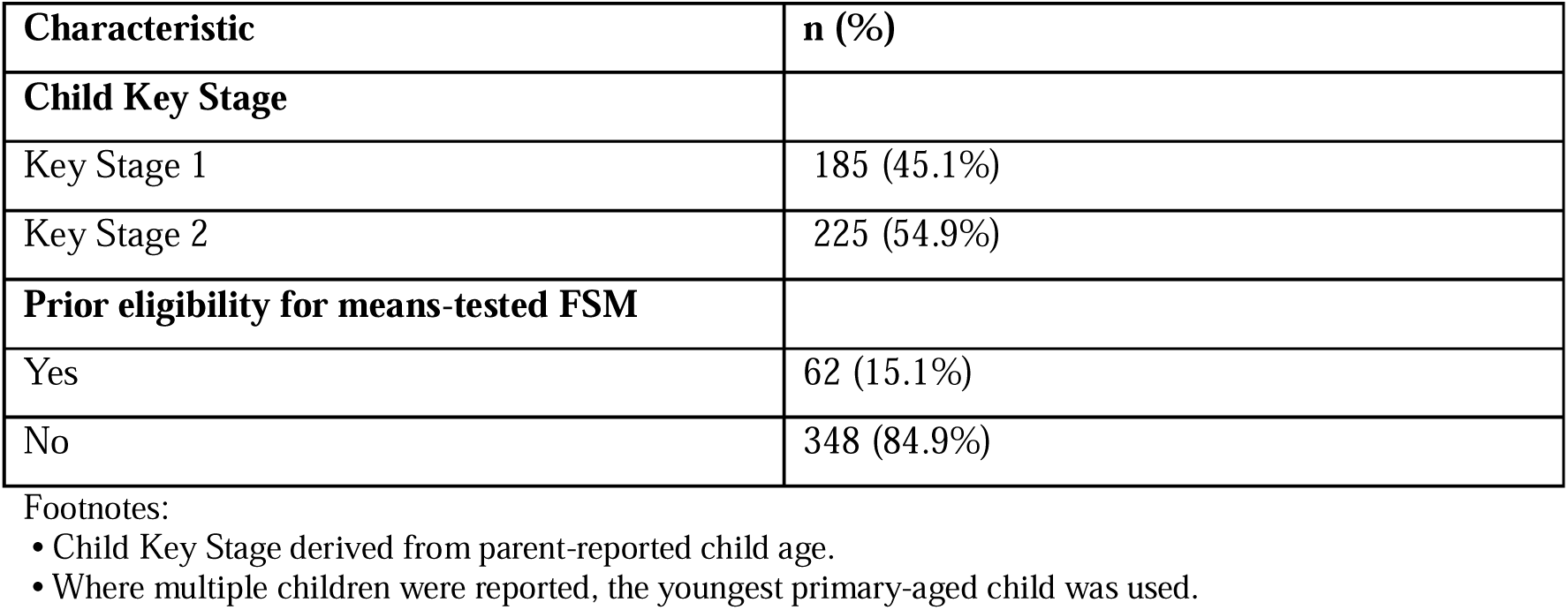
Demographic information.

Parents frequently expressed mixed feelings about UFSM. While many supported the policy in principle, they raised significant concerns about the quality, nutritional value and quantity of the meals provided. We identified two commonly occurring themes that reflect parent’s priorities and decision-making around UFSM. These are presented below. Full themes table can be found in supplementary (S4).

### Theme one – The food

A recurring issue across responses was that portion sizes were considered too small, particularly for older or more active children. Several parents noted that their children often returned home hungry, suggesting that meals did not provide sufficient sustenance throughout the school day:

*“The food portions are insufficient, my child tells me he gets 1/3 of the size we give him at home. Different choices and sides to fill out the meal would be much more beneficial.”* (Parent 229)

*“I think free school meals initiative is great, but I do strongly feel that the portion sizes need to be looked at, as often my child comes home hungry as he says he wasn’t given enough food, and sometimes the canteen runs out of food so don’t have a full meal.”* (Parent 285)

The quality and nutritional value of school meals was also noted significantly. Parents described the food as bland, overly processed or of poor standards:

*“Quality of food is poor, little taste, small portions for the older children and lack of cleanliness – hair is frequently found in food. Overall school meals are still a disgrace.”* (Parent 317)

*“Better quality of meat, less processed rubbish. Uniformed cubed chicken does not look appetising and it tastes rubbery.”* (Parent 405)

Parents also raised concerns about the implications for children’s health, expressing their children are being served ultra-processed foods regularly and want to see healthier meals provided:

*“Excellent scheme, fully support it happening. However, I am concerned about the health implications of routinely eating ultra-processed foods in early childhood.”* (Parent 312)

*“My main concern is that they often include ultra-processed foods. As a single parent on a tight budget, it would be great to take up free school meals, but it’s important to me that my son has a healthy lunch.”* (Parent 351)

*“Nutritional quality of the food is very poor. The food they seem to offer is very processed and especially high in salt.”* (Parent 345)

Although many parents recognised the benefits of UFSM and welcome the provision, concerns around portion sizes, quality and nutritional value of the meals provided are concerning.

### Theme two: Factors influencing uptake

Parents who chose to provide their children with a packed lunch, either regularly or occasionally, often described doing so as they could better control their children’s diet, health and wellbeing. For many, providing food from home ensured that their child received meals that were both appealing and aligned with family values around health and nutrition.

Some parents linked their decision to their child’s dietary goals or health motivations, particularly when children were involved in sports or personal fitness:

*“My son has only recently switched to packed lunches as he really wants to get fitter for his sporting activities and he doesn’t feel school meals offer this opportunity.”* (Parent 15)

Others were influenced by concerns about portion size and satisfaction, describing school meals as insufficient for older children:

*“As my child doesn’t like some of the meals and also there is not enough food on the meal trays for older children like my own child…they only have about 5 chips and 3 nuggets and a spoon of peas. I don’t think that is enough for a year 5 or 6 pupil.”* (Parent 110)

Food preferences and peer influences also played a role, with some parents noting that their child’s choices reflected social patterns as well as taste:

*“My daughter isn’t a massive meat eater, she very often has the veggie option. But she isn’t keen on the roast dinner veggie option on a Wednesday. She is also influenced by most of her friendship group having packed lunch on a Wednesday.”* (Parent 155)

Finally, several parents cited trust and safety concerns, particularly where allergies were present. For these families, sending a packed lunch was viewed as a necessary safeguard:

*“My son has allergies, so I got the chance to take a look at all the ingredients in the school kitchen and everything was in a jar! Pasta sauce, pizzas were already prepared. Nothing fresh! My son didn’t respond well to even the allergen free meals. There was sugar in everything.”* (Parent 320)

Overall, these responses show that decisions to opt out of universal free school meals are influenced by a mix of health beliefs, food preferences, social contexts, and trust in meal preparation. For many families, packed lunches offer a sense of reassurance and agency, factors that remain significant even when meals are available at no cost.

Alongside the open-ended comments above, parents completed several closed-ended items further exploring their views on UFSM and its impact on their children and household.

Parents held mixed perceptions on nutritional quality, portion size and meal satisfaction, as well as social and emotional impacts. Findings are presented in table 2. The table presents the percentage of parents who agreed, were neutral, or disagreed with each closed-ended survey item, grouped by theme (n = 410).

**Table 2a:**
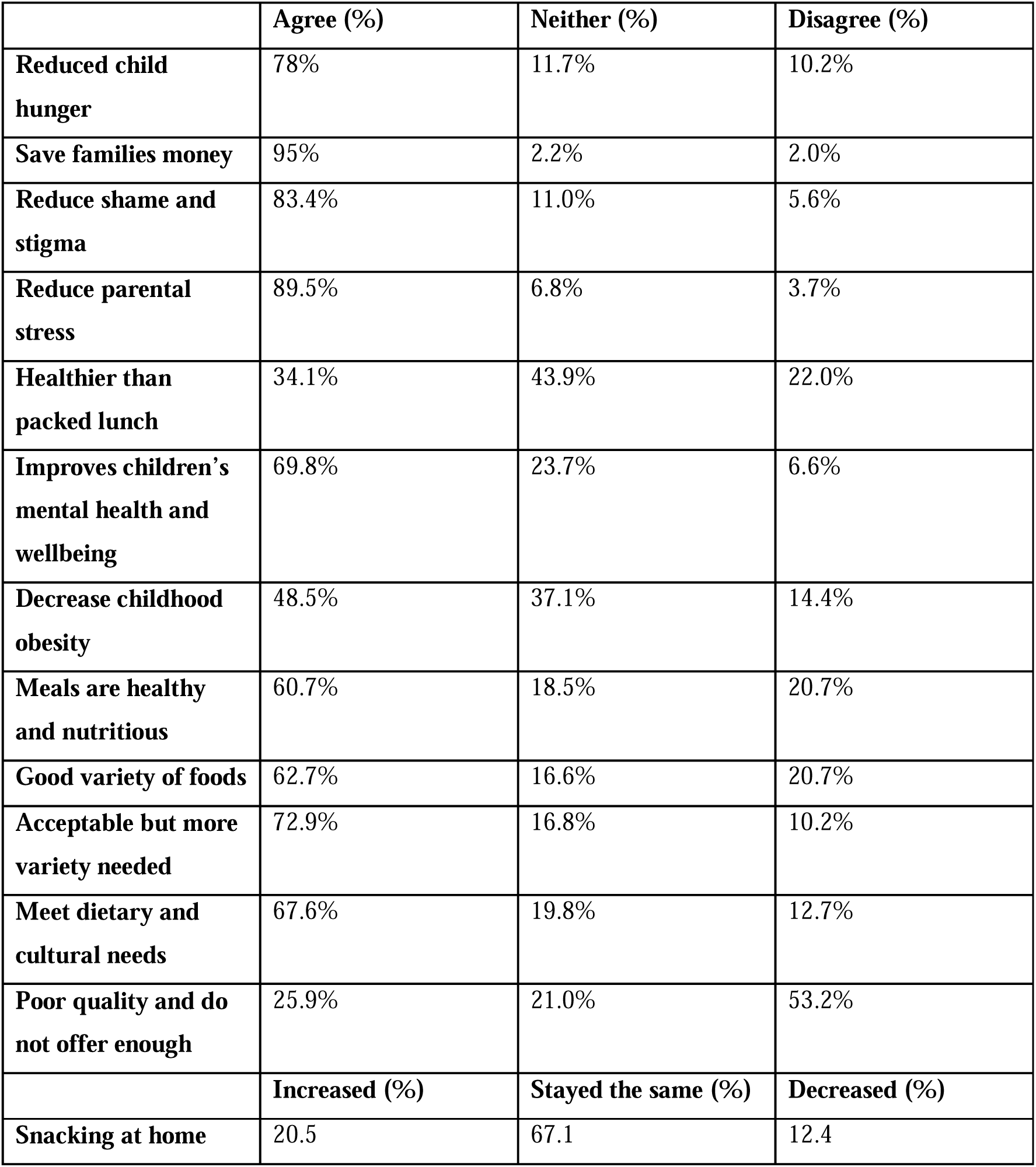
Closed ended responses from parent survey.

### Summary of overall findings

As seen, UFSM was widely welcomed for promoting fairness, reducing stigma and ensuring access to at least one nutritious meal a day. Children and parents both emphasised positive emotional and social effects of shared mealtimes, describing lunchtime as an opportunity for inclusion, conversation and play. However, persistent concerns about meal quality, variety and portion size limited uptake, with parent also expressing a need for more flexible dietary options. These findings suggest that while UFSM is welcomed, it’s long-term success and improving uptake is dependent on improvements in menu diversity and better-quality food.

### School infrastructure and lunchtime organisation

Observations made by the research team during school visits saw notable variations in schools’ lunchtime infrastructure and organisation, which appeared to shape children’s lunchtime experiences. In one school, the head teacher explained that an additional dining hall was being constructed to accommodate the increased number of children accessing hot meals following the introduction of UFSM. At the time of data collection, limited dining space saw some children having to eat their meals in classrooms.

Across several schools, lunch was organised in phased sittings because of limited space, with younger children typically eating first and older children later. This arrangement often reduced the time available for older children to eat and play and, in some cases limited opportunities to sit with peers of their choosing. These organisational constraints led to some children reporting feelings of unfairness and reduced social and play time.

In contrast, focus groups conducted in schools with sufficient dining space and more flexible lunchtime arrangements reported more favourable lunchtime experiences. Children who had strong relationships with catering staff also expressed more positive views, including more enjoyment of the meals themselves as well as the wider dining environment.

## Discussion

This study explored the perspectives of children and parents on the implementation and impact of UFSM in Wales. Findings demonstrate that both children and parents welcome the provision, however significant improvements are needed for the provision to meet its full potential. These findings underline that the effectiveness of UFSM depends not only on the provision itself but on the quality, organisation, and social experience of mealtimes within schools.

### The Food Experience: Quality, Variety and Sustenance

Both children and parents described food quality, portion sizes and variety as key determinants of whether UFSM was viewed positively. Many children valued trying new foods or eating a hot meal with friends, while others expressed dissatisfaction with repetitive menus. Meals were also perceived as bland or insufficient to meet their needs. Parents echoed these concerns, often linking portion sizes and food quality to whether children came home hungry or opted to take a packed lunch from home instead.

Previous UK research has shown perceptions of menu variety, food processing and portion size can act as barriers to uptake of school meals are key factors in influencing up-take [28].

Parents’ concerns about ultra-processed food and nutritional quality also mirror growing public health debates about the long-term implications of dietary quality in early childhood [29]. These findings highlight the importance of improving school food standards, variety and portion sizes to meet the intended equity and health objectives of the provision outlined by the Welsh Government [11].

### The Social Value of Lunchtime

Children’s reflections highlighted that the benefits of UFSM extend beyond nutrition. Lunchtime was described as a valued time to connect with peers, build a sense of belonging and play, with children frequently mentioning wanting time with friends at lunchtime. These findings align with work in Norway, which found shared school meals improved peer-relationships [17].

Findings from this study also support previous research from the United States, where the introduction of UFSM was linked to improved social climate in schools and supports children’s social development and emotional wellbeing [18]. In this study, children described lunchtime as an opportunity for social interaction and play, highlighting its role in for relationship building, connectiveness and a sense of belonging. Such interactions are fundamental to children’s social learning, helping to build empathy, cooperation and communication skills that extend beyond the dining hall [30]. When mealtimes allowed children to sit together and engage freely, they reported greater enjoyment, reinforcing the view that shared meals contribute to a positive psychosocial environment [17,18]. However, this study found that logistical constraints, such as limited dining space and fixed seating arrangements restricted children’s ability to sit with friends, promoting feelings of unfairness. This reinforces that the social climate of lunch time is as critical to children’s development as the meals itself, shaping how UFSM is experienced.

### Parental Decision-Making and Uptake

Parents decisions to provide their children with a packed lunch, even when UFSM were available, reflected the importance of parents having agency over their children’s health and wellbeing. Many parents viewed packed lunches as a means of ensuring their children consumed foods that aligned with their preferences and nutritional expectations, particularly when they perceived school meals to be overly processed or lacking in variety. As mentioned above, similar patterns were reported by Sahota et al., [28], where parents confidence in food quality and freshness was influenced uptake.

Concerns around portion sizes and meal satisfaction were also important to decision making. Several parents noted their children came home hungry, or that older children were not served enough. For some parents, the perception that meals were insufficient or unbalanced diminished trust in the policy’s ability to meet children’s nutritional needs.

At the same time, many parents acknowledged the significant benefit to their household.

Responses to the closed ended survey questions indicated that the UFSM policy helped reduce financial strain, a valuable source of cost savings during a period of rising living expenses. Parents also reported that UFSM reduced morning stress and time pressures, as it alleviated the need to buy and prepare lunches daily. These findings show that whilst some families continue to opt out for reasons relating to food quality and dietary control, others view the policy as a practical investment which supports family wellbeing.

### Strengths and Limitations

A key strength of this study is its multi-perspective, mixed-method design, which allowed for an in-depth exploration of how UFSM is experienced by both children and parents across diverse geographical and socio-economic areas in Wales. The use of focus groups with children provided first hand insights into children’s everyday experiences of school meal capturing authentic voices that are often underrepresented in policy discussions.

Complementing this, the parent survey combined open-and closed-ended questions, offering both quantitative context and qualitative depth to understand family-level decision-making and perceptions of UFSM.

However, several limitations should be acknowledged. The study included a limited number of schools (n = 8), and participation was based on convenience sampling, meaning that findings may not represent all schools or regions in Wales. Children were recruited from Year 6 only; thus, younger learners were not represented. Finally, as data was collected during the early stages of UFSM implementation, perspectives may evolve as the policy becomes more embedded and operational challenges are addressed.

### Implications for Policy and Practice

The findings from this study highlight several areas for consideration to maximise the impact of UFSM in Wales. First, consistency in food quality, portion sizes and menu variety is essential to ensure meals are both nutritionally valuable and appealing to children. In turn, encouraging improved uptake across Welsh primary schools. Additionally, investment in fresh, less processed meals and the inclusion of more varied meal options could help enhance the acceptability of meals for all children as well as build greater confidence among parents in the quality and nutritional value of the food provided.

Second, schools should recognise lunchtime as a social and emotional environment as well as a nutritional one. Providing sufficient time, space and flexibility for children to sit and enjoy time spent with peers is vital for emotional and social wellbeing. Our findings showed that when children were able to sit with friends, make plans and transition easily from eating to play, lunchtime became one of the most enjoyable parts of their school day. When schools had rigid seating structures or shortened lunchtimes, children expressed feelings of unfairness. Ensuring that lunchtime routines allow adequate time for eating and play can therefore enhance both the social and emotional benefits of UFSM.

## Conclusion

This study explored children’s and parents’ experiences of UFSM policy in Wales. There is support for promoting inclusion, access to food whilst also easing financial and practical pressures for families. However, significant concerns were raised around the quality of food, portion sizes and variety offered, thus limiting its benefits. For children lunchtime is not just a time to eat but also about social connection and play contributing to enjoyment and a sense of belonging. Parents valued the policy’s financial relief but expressed concerns about nutritional value, food processing, variety, all of which shaped their trust and contributed to uptake decisions. With continued investment in portion sizes, menu variety, and consistent provision of fruit and vegetables, UFSM could make a meaningful contribution to healthier dietary patterns and long-term health outcomes for children in Wales. UFSM policy may benefit from greater service user consultation to ensure children’s and parents’ perspectives inform ongoing policy refinement. Recognising lunchtime as both a nutritional and social opportunity will further strengthen the benefits of UFSM and support positive mealtime environments.

## Acknowledgements Funding

Funding was provided by Swansea University as a PhD studentship and supported by the Centre for Population Health. The funders had no role in study design, data collection and analysis, decision to publish, or preparation of the manuscript.

## Author Contributions

AL is responsible for the overall content of as the guarantor, led the study and was responsible for collecting and analysing the data and preparing the original draft. MJ provided supervision, collected data and editing.

SB provided supervision and editing.

## Patient and public involvement

Patients and/or the public were not involved in the design, conduct, reporting or dissemination plans of this research.

## Data availability

Due to the nature of qualitative data, transcripts cannot be shared publicly. Aggregated survey data may be available upon reasonable request.

## Ethics statement

This study involves human participants and HAPPEN has been granted ethical approval by Swansea Universities School of Medicine (ethics board ref: 7933).

