## Supplementary 1 for "Children’s and Parents’ Perspectives on Universal Free School Meals in Wales: A Mixed Methods Study on Health, Wellbeing and Barriers to Uptake"

**Focus group Questions (Children)**

1. What do you think about free school meals for everyone? Do you think school meals should be free for everyone? Why is this?
2. Can you describe a meal you've had at school that you really enjoyed? What made it so good?
3. Can you describe a meal you've had at school that you didn’t enjoy? What didn’t you like about it?

4. If you could create your own school lunch menu, what would it include?
5. How do you decide who to sit with during lunchtime at school? Do you sit by the same person/people or different?
6. What sorts of things do you enjoy talking about or doing with your friends while you eat school lunch?
7. How do you think having a school meal affects other parts of your day? i.e., schoolwork, energy etc.
8. Do you eat school meals every day? Bring packed lunch on some days? – why is this?
9. How do you feel after eating a meal at school compared to when you eat a packed lunch?
10. Have you ever tried a new food at school that you ended up really liking? What was it, and what made you decide to try it?
11. Is there anything you would tell us about Free school meals that we haven’t covered yet?
