## Supplementary 2 for "Children’s and Parents’ Perspectives on Universal Free School Meals in Wales: A Mixed Methods Study on Health, Wellbeing and Barriers to Uptake"

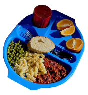

### Your Thoughts on School Meals

Are you a parent or guardian of a primary school child in Wales? We'd love to hear your thoughts on universal free school meals. Your insights will help us better understand the impact of this policy on children and families.

This survey will take 5-10 minutes to complete.

For more information about the study and how your responses will be used, please visit: [https://happen-wales.co.uk/upfsmthoughts/](https://happen-wales.co.uk/upfsmthoughts/)

Thank you!

* Required

#### Consent

1. I have read and understood the information about the study *

- ◯ Yes
- ◯ No

2. I consent to taking part in this study *

- ◯ Yes
- ◯ No

#### Survey

3. **How much do you agree or disagree with the following statements regarding universal free school meals?** *

| | Strongly Disagree | Disagree | Neither Agree or Disagree | Agree | Strongly Agree |
|---|---|---|---|---|---|
| Free School Meals reduce child hunger | ○ | ○ | ○ | ○ | ○ |
| School meals save families money | ○ | ○ | ○ | ○ | ○ |
| They reduce shame and stigma in the lunch room | ○ | ○ | ○ | ○ | ○ |
| They provide meals that are healthier than meals brought from home | ○ | ○ | ○ | ○ | ○ |
| School meals can improve children’s mental health and wellbeing | ○ | ○ | ○ | ○ | ○ |
| They can decrease childhood obesity | ○ | ○ | ○ | ○ | ○ |
| They can help reduce parental stress | ○ | ○ | ○ | ○ | ○ |

4. **How much do you agree or disagree with the following statements regarding the quality and variety of universal free school meals?** *

| | Strongly Disagree | Disagree | Neither Agree or Disagree | Agree | Strongly Agree |
|---|---|---|---|---|---|
| Meals are healthy and nutritious, meeting all dietary standards | ◯ | ◯ | ◯ | ◯ | ◯ |
| Meals include a good variety of foods, catering to diverse tastes and preferences | ◯ | ◯ | ◯ | ◯ | ◯ |
| Meals are acceptable, but more variety could be introduced to keep them interesting | ◯ | ◯ | ◯ | ◯ | ◯ |
| Meals meet my child's dietary and cultural needs | ◯ | ◯ | ◯ | ◯ | ◯ |
| Meals are of poor quality and do not offer enough variety | ◯ | ◯ | ◯ | ◯ | ◯ |

5. Since the introduction of universal free school meals, has your child's snacking at home: *

◯ Increased

◯ Decreased

◯ Stayed the same

6. **How often does your child bring a packed lunch to school?** *

◯ Never

◯ One to two days a week

◯ Three to four days a week

◯ Always - they do not have a school meal

7. What influences the amount of days your child brings a packed lunch to school? *

8. **Do you think the school meals program has affected your child's understanding or awareness of healthy eating?** *

- ◯ Yes
- ◯ No

9. **Thinking about the roll-out of free school meals for everyone, which of the following statements have you experienced regarding your child's physical health?** *

Select all that apply

- ☐ My child has increased energy after having a school meal
- ☐ My child sleeps better after having a school meal
- ☐ My child has less illnesses since free school meals
- ☐ My child's overall physical health has improved
- ☐ I have not seen any changes
- ☐ Other

10. **Thinking about the roll-out of free school meals for everyone, which of the following statements are true regarding your child's emotional health?** *

Select all that apply

- ☐ My child's mood has improved.
- ☐ My child's behaviour at home and/or at school has improved.
- ☐ My child has improved concentration/focus.
- ☐ My child's overall mental health and wellbeing has improved.
- ☐ I have not seen any changes.
- ☐ Other

11. **Are there any changes or improvements you would like to see made to school meals?** *

12. **Do you have any other feedback/concerns/general comments relating to universal free school meals?** *

13. How old is your child? *

14. What school does your child attend? *

15. Prior to the roll-out of universal free school meals, my household was entitled to means-tested free school meals *

- Yes
- No

This content is neither created nor endorsed by Microsoft. The data you submit will be sent to the form owner.

Microsoft Forms
