## Supplementary 3 for "Children’s and Parents’ Perspectives on Universal Free School Meals in Wales: A Mixed Methods Study on Health, Wellbeing and Barriers to Uptake"

| **Themes** | **Sub-themes** | **Quotes** |
| --- | --- | --- |
| **Theme One: The Food Experience** | **Taste, enjoyment, and appeal**  **Food freshness and temperature     Portion size and feeling satisfied**  **Variety, repetition, and choice.** | *“The sausages taste weird, like they’ve been there a while.”* **(Child, Focus group 4)** *“They’re quite bad now… the stuff doesn’t taste nice.”* **(Child 2, Focus group 5)** *“I like the fish fingers because I’d never tried them before and it caught my attention. It was really good.”* **(Child 4, Focus group 5)** *“Brunch or the chicken wrap because I love the chicken… the sausage is cooked really nice!”* **(Child 7, School 4)** *“All the dinners are really nice, I always have a school dinner because I think they are better than a packed lunch.”* **(Child 7, Focus group 4)**  *“Sometimes it’s gone cold by the time you get it.”* **(Child 5, Focus group 2)** *“The sausages taste weird, like they’ve been there a while.”* **(Child 8, Focus group 4)**  *“On Friday when they do fish, they only give you like two fish fingers.”* **(Child 3, Focus group 2)** *“For the little ones yes, but for us no, because we are growing.”* **(Child 7, Focus group 4).** *“We get enough food, sometimes too much so we are really full and don’t want to move.”* **(Child 1, Focus group 8).**   *“say theres like chicken or roast dinner, like id have to have the veggie one instead.”* **(Child 3, School 5)** *“They’re quite bad now, especially for vegetarians. There isn’t enough choice.”* **(Child 2, Focus group 5)** *“I think there should be more variety… it just goes through that routine and I don’t particularly like the options.”* **(Child 5, School 5)** *“I like a mix, some days I choose cake and some days I choose the fruit. There is usually two things to choose from.”* **(Child 5, Focus group 7).** |
| **Theme two: The Social Value of Lunchtime** | **Sitting with friends and social inclusion**  **Playtime as a central priority**  **Time pressure and trade-offs between eating** **and play** | *“It’s not fair, because you want to sit by your friends. Like I sit by all the boys and it’s not fair.”* **(Child 5, Focus group 3)** *“We normally just go in and hope we can sit by our friends, because we don’t always get to choose.”* **(Child 4, Focus group 3)** *“It is better now because it used to be pack lunches on one table and dinners on another, so we didn’t get to sit with your friends if they were all packed lunch and you were dinners.”* **(Child 5, Focus group 3)**  *“Then as soon as we’re done you could just bolt it for the field.”* ***(Child 7, Focus group 3)*** *“So we could play!”* **(Child 3, Focus group 3)** *“The younger ones go for food first so we go out to play and then have food and if we finish our food quickly, we can go back out to play.”* **(Child 2, Focus group 8)** *“The dinner lady makes the pudding, she’s nice, but it takes time to make the pudding so people rush their food and then waste it because they want to go out to play.”* **(Child 3, Focus group 4)** *“We make plans about what we are going to do after school or games we are going to play on.”* **(Child 5, Focus group 3)**  *“Yeah then we go last and only have about 15–20 minutes to eat our food and then we have about 10 minutes to play after.”* **(Child 8, Focus group 4)** *“People rush their food and then waste it because they want to go out to play.”* **(Child 3, Focus group 4)** |
| **Theme three: Fuel for Learning and Feeling Good** | **Food for concentration and learning**  **Mood and energy** | *“When I have had a good meal, I won’t be thinking I want food because I’m hungry. I can think about the work.”* **(Child 1, Focus group 4)**  *“If we don’t have enough food we aren’t full so we are hungry and can’t think.”* **(Child 2, Focus group 4)**  *“yes because if your hungry your focusing on that”* **(Child 4, Focus group 1)**  *“ I feel tempted to eat stuff from my lunch in the day but if I was hot dinners then obviously I wouldn’t be able to.”* **(Child 3, Focus group 1)**  *“If it looks nice, I will have a good day, but if it doesn’t look nice then I don’t want to eat it and I’ll have a bad day for the rest of the day.”* **(Child 2, Focus group 6)**  *“I think the portions are sometimes too much so its hard to have energy after eating it.”* **(Child 4, Focus group 5)** *“School meals actually help me have energy through the day.”* **(Child 5, Focus group 2)** *“It cheers you up so your less grumpy.”* **(Child 2, Focus group 1)** |
