## Supplementary 4 for "Children’s and Parents’ Perspectives on Universal Free School Meals in Wales: A Mixed Methods Study on Health, Wellbeing and Barriers to Uptake"

| **Theme** | **Subthemes** | **Associated Quotes** |
| --- | --- | --- |
| **Theme 1: The food** | **Perceived poor food quality**  **Limited variety and repetitive menus**  **Ultra-processed and nutritional concerns**  **Inadequate portion sizes** | *“Quality of food is poor, little taste, small portions for the older children.”* *(Parent 317)*  *“The quality of ingredients need to he improved, more fresh ingredients and less processed food - it's the reason my children don't really like them.”* **(Parent 9)**  *“Quality of food is poor, little taste, small portions for the older children and lack of cleanliness – hair is frequently found in food. Overall school meals are still a disgrace.”* (Parent 317)  *“Better quality of meat, less processed rubbish. Uniformed cubed chicken does not look appetising and it tastes rubbery.”* (Parent 405)  *“More variety/options.”* **(Parent 1)**  *“Certain days there are no suitable meal options for my child.”* **(Parent 3)**  *“Some of the options on the menu are quite frankly bizarre. One example… naan with sweet and sour chicken and salad of cucumbers/peppers.”* ***(Parent 13)***  *“Excellent scheme, fully support it happening. However, I am concerned about the health implications of routinely eating ultra-processed foods in early childhood.”* (Parent 312)  *“My main concern is that they often include ultra-processed foods. As a single parent on a tight budget, it would be great to take up free school meals, but it's important to me that my son has a healthy lunch.”* (Parent 351)  *“Nutritional quality of the food is very poor. The food they seem to offer is very processed and especially high in salt.”* **(Parent 345)**  *“I am concerned about the health implications of routinely eating ultra-processed foods in early childhood.”* **(Parent 312)**  *“The food they seem to offer is very processed and especially high in salt.”* **(Parent 345)**  *“The food portions are insufficient, my child tells me he gets 1/3 of the size we give him at home. Different choices and sides to fill out the meal would be much more beneficial.”* (Parent 229)  *“I think free school meals initiative is great, but I do strongly feel that the portion sizes need to be looked at, as often my child comes home hungry as he says he wasn't given enough food, and sometimes the canteen runs out of food so don’t have a full meal.”* (Parent 285)  *“There is not enough food for the children. Portion sizes are terrible… same size portion for a 5 year old as a 10 year old… food runs out.”* **(Parent 7)**  *“The food portions are insufficient, my child tells me he gets 1/3 of the size we give him at home.”* **(Parent 229)** |
| **Theme Two: Factors Influencing Uptake** | **Control over diet and health**  **Trust, safety and allergies**  **Peer influence**  **Menu and preference** | *“As a single parent on a tight budget, it would be great to take up free school meals, but it's important to me that my son has a healthy lunch.”* **(Parent 351)**  *“There is also no way for parents to know the volume of food offered or how much is actually eaten, so it is difficult to get an understanding of what impact this would have on my child's diet overall.”* **(Parent 340)**  *“My son has only recently switched to packed lunches as he really wants to get fitter for his sporting activities and he doesn’t feel school meals offer this opportunity.”* **(Parent 15)**  *“More variety for those with dietary requirements for medical reasons (eg gluten free/diabetic etc)”* **(Parent 128)**  *“More choices for children with allergies. My eldest has a tomato allergy and lots of the meals include tomato based sauces.”* **(Parent 95)**  *“More support for schools in relation to dietary requirements and allergies.”* **(Parent 148)**  *“My son has allergies, so I got the chance to take a look at all the ingredients in the school kitchen and everything was in a jar! Pasta sauce, pizzas were already prepared. Nothing fresh! My son didn’t respond well to even the allergen free meals. There was sugar in everything.”* **(Parent 320)**  *“she isn’t keen on the roast dinner veggie option on a Wednesday. She is also influenced by most of her friendship group having packed lunch on a Wednesday.”* (Parent 155)  *“Who they want to sit next to at lunch time (school dinners sit separately to packed lunches).”* **(Parent 11)**  *“Where they get to sit. If friends are having packed lunch, they want one so they can sit next to that friend.”* **(Parent 178)**  *“As my child doesn't like some of the meals and also there is not enough food on the meal trays for older children like my own child … they only have about 5 chips and 3 nuggets and a spoon of peas. I don't think that is enough for a year 5 or 6 pupil.”* (Parent 110)  *“Child only has school lunch Friday as doesn’t like the set menu for other 4 days no options”* **(Parent 73)**  *“My child prefers to have school dinners and enjoys getting to choose the meals she has each day.”* **(Parent 94)**  *“Its my childs preference, we limit it to two days due to cost and time preparing.”* **(Parent 83)** |
